# Medical students’ perception of resilience and of an innovative curriculum-based resilience skills building course: a participant-focused qualitative analysis

**DOI:** 10.1101/2023.01.03.23284139

**Authors:** Bhavana Nair, Farah Otaki, Avantika Fiza Nair, Samuel B. Ho

## Abstract

**Background:** Medicine is one of the most demanding academic fields with an extensive curriculum that entails plenty of potential stressors. There is sufficient evidence that medical students are more prone to psychological distress when compared to their peer group of other disciplines. Despite the established need to prioritize resilience skills building within the medical curriculum, very few medical programmes in the Middle East and North Africa region (MENA) proactively empower the students to help themselves in sustaining their mental health. The purpose of the current study is to explore the perception of medical students in Dubai, United Arab Emirates (UAE) regarding their understanding of, and personal experience with building resilience, and their engagement with the content of an innovative curriculum-based resilience skills building course, designed in alignment with the constructivism theory of education.

**Method:** The current study utilized a qualitative phenomenological research design. The curriculum-based resilience skills building course, that was investigated as part of this study, is offered at a medical school in Dubai, UAE. A total of 37 students submitted reflective essays about resilience building, in general, and the respective course, in specific. The collected data was inductively analysed following a six-step framework.

**Findings:** The qualitative analysis generated three interlinked themes, namely: Awareness, Application, and Appraisal.

**Conclusion:** This study showed that integrating a resilience skills building course into medical curricula is likely to be positively appraised by the students, where it raises their level of awareness and likelihood of proactively applying the learned concepts in their daily lives. This is especially true when the course is anchored in constructivism experiential learning theory and designed to foster self-directed learning.

## Introduction

Medicine is one of the most demanding academic fields with an extensive curriculum that entails plenty of potential stressors. These include but are not limited to concerns related to adjusting to the medical environment, handling difficult patients, attaining work-life balance, sleep deprivation, feeling overwhelmed due to information overload, and financial pressures. These challenges naturally put students at a greater risk of stress and anxiety [1].

Job-related distress or “burnout” in medical trainees and healthcare professionals is an increasingly recognized problem worldwide. Despite the fact that medical students can avail health-related services easily, they often are less likely to access treatment in comparison with the general population [2, 3], leading to a higher incidence rate among them to developing and experiencing feelings that are unpleasant which are typically referred to as psychological distress [4] [5].

There is sufficient evidence that medical students are more prone to psychological distress when compared to their peer group of other disciplines [6]. In addition to that, the COVID-19 pandemic witnessed a surge in susceptibility to deteriorating quarantine-related mental health among the public [7] especially among medical students whose academic routine got significantly affected. This was associated with an increase of anxiety, depression, and irritability [8-10]. The tendency to overconsume information related to the pandemic exaggerated the symptoms of stress and anxiety in a section of people who were already pre-disposed to the condition, thereby spiking their anxiety to even more heightened levels [10]. Hence, it is not surprising that burnout, among practicing doctors and medical trainees, was increasingly reported upon [11]. In a sense, this global crisis further emphasized the importance of building resilience skills, among students, highlighting the need for them to be equipped with adaptive skills to handle the stressors of a challenging learning environment. People react to stress in different ways. The capability to adjust to adverse challenges and events is often referred to as Resilience [12]. There seems to be an inversely proportional relationship between resilience, and susceptibility to pathological ways of coping with adverse conditions [13].

The transactional stress model theory proposes that an individual’s capacity to cope and adjust to challenges and problems is a consequence of transactions that occur between the respective individual and their environment. It highlights the significance of one’s understanding of and behaviour towards the subject matter in their handling of stress [14]. As such, the individual’s evaluation and appraisal of a stressful situation influences the experience of the stress, and the current generation of students think very differently from earlier generations with regard to their mental health requirements [15]. As the student population in higher education is becoming increasingly more overwhelmed with external as well as internal stressors, a holistic approach to well-being [e.g., 4M-Model of Mindfulness, Movement, Meaning, and Moderator [15]] is needed which in turn is expected to foster self-directed learning [16]. Self-directed learning involves forethought and planning, analysis, and implementation of clear goals as well as reflection of performance which requires self-control and initiative from students [17].

This was witnessed during the lockdown and online learning during the COVID-19 pandemic as university students were forced to use more self-regulated processes to strategize and to achieve their goals due to the isolation [18-21].

There are several medical programs, offered in North America, that have a curricular component that aims at developing the learners’ resilience skills [22]. Most of those programs are designed in a way to foster self-directed learning, assuming the learners are adults who engage in the learning activity for its inherent satisfaction rather than for some separable consequence, and who had integrated from previous learning experiences substantial amount of knowledge [23, 24]. These learners are assumed to have the ability to perceive and use relational similarity between two situations. As such, in alignment with Kolb’s interpretation of experiential learning[25]. Experiential learning: Experience as the source of learning and development. FT press.), the learning becomes cyclically composed of the following phases: concrete experience, reflective observation, abstract conceptualization, and active experimentation[26]. It will be worthwhile to further reinforce such resilience skills building courses by anchoring them in the constructivism experiential learning theory which views learning as a process of active adaption [27, 28].This theory presents learning as an experience and participation [23, 27].

Despite the established need to prioritize resilience skills building within the medical curriculum, very few medical programmes (if any) in the Middle East and North Africa region (MENA) proactively empower the students to help themselves in sustaining their mental health [29]. Accordingly, the purpose of the current study is to explore the perception of medical students in Dubai, United Arab Emirates (UAE) regarding their understanding of, and personal experience with building resilience, and their engagement with the content of an innovative curriculum-based resilience skills building course, which is designed in alignment with the constructivism theory of education.

## Methods

### Context of the study

This study took place at the Mohammed Bin Rashid University of Medicine and Health Sciences (MBRU), Dubai, UAE in the College of Medicine (CoM). One of the degree programs offered at the respective college is a six-year full-time Bachelor of Medicine and Bachelor of Surgery program (MBBS). The course of study is divided into three phases as depicted below (Fig. 1).

**Figure 1.**
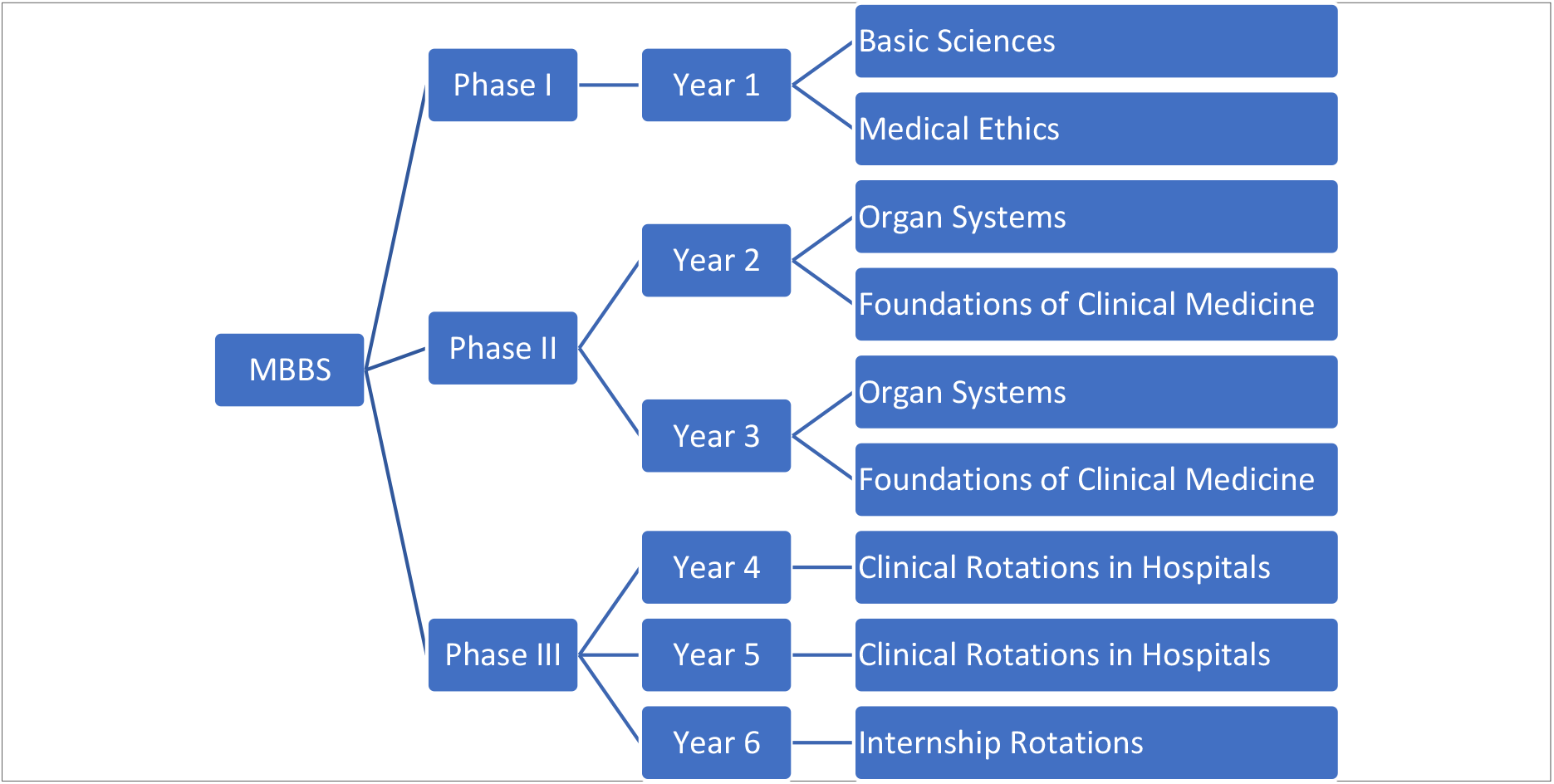
Outline of Structure of MBBS

### Description of the innovative curriculum-based resilience skills building course

This course (Appendix: Course Guide) consists of 6 hours of instruction on building skills for resilience during the first clinical year of the MBBS (i.e., Year 4). In the respective year, the students are rotating in clinical placements for four days a week and are attending classes on campus once a week (labelled as “MBRU Day”). During the “MBRU Day”, students are offered structured, curricular training across various longitudinal themes, including but not limited to the resilience skills building course. The overall objective of this course is to raise awareness about the challenge of stress in the medical students’ trajectory and the clinical workplace, and to provide tools for understanding, developing, and deploying resilience skills. By the end of the course, the students are expected to be able to:

- Understand the concept of work-place stress and burnout
- Understand the concept of resilience
- Develop an understanding of the wide variety of tools and techniques that are available to develop and activate resilience
- Comprehend basics of Cognitive Behavioural Therapy
- Demonstrate an understanding of healthy coping strategies
- Understand mindfulness and meditation practices
- Develop knowledge about the skills related to emotional intelligence
- Understand the basic characteristics of mental toughness
- Describe the practice and benefits of gratitude
- Raise self-awareness and perceived self-efficacy
- Enhance one’s capacity for emotional self-regulation

The course is designed in a way that inspires and empowers adult learners [28] who are assumed to be self-directed and are intrinsically motivated[27].The postulation is that these learners tend to exercise analogical reasoning in learning and practice. They have gone through diverse learning experiences in basic and clinical sciences. As such, these learners have retained a substantial knowledge base which constitute an increasing resource for learning, and forms mental models which drive their attitudes and behaviours. Through the course and their engagement with the course content, based on the constructivism theory of experiential education [24], the students are encouraged to identify gaps in their own mental models and to adapt them based on their active participation in learning experiences. In alignment with Kolb’s experiential learning theory, reflection and reflexivity are fostered through the supervision of and continuous flow of feedback from skilled mentors, who are experts in the subject matter [23, 27]. Moreover, students are asked to maintain a daily journal of reflections, where they document what surfaces for them during the session. Also, at the end of each session, the students are asked through an online survey to pinpoint one or two main take-home messages as well as the actions they intend to take to proactively build their resilience. As such, in between the weekly course sessions, the learners get to actively experiment with the application of the acquired knowledge and skills.

This (pass or fail) longitudinal course has three student performance assessment components: attendance requirement, Objective Structured Clinical Examination (OSCE), and reflective essays. Enrolled students are expected to attend all the course classes. Students who miss more than 20% of the class sessions are automatically dropped-out from the course. Students are required to come on time to each session. The OSCE component of this course is factored into the end of the academic year assessment, where students are practically tested (through simulations) on various skills including those related to resilience. As for the essay, the students are required to submit a 500 words essay reflecting on their learning experience as part of this course.

### Research design

The current study utilized a qualitative phenomenological research design [30].This interpretive participant-focused design enabled the researchers to uncover what the phenomenon of building resilience, and the learning experience integral to the respective longitudinal course, mean to the participants. This approach required the researchers to thoroughly understand the participants’ descriptions of their lived experiences [31]. The participants had complete autonomy to choose whether, or not, to participate. This study was approved by the Institutional Review Board of MBRU (MBRU-IRB-2019-021).

### Data collection

The data collection for the current study was integral to the respective resilience skills building course. The students’ submissions for the reflective essay requirement (which allowed for substantial reflectivity and reflexivity) constituted the dataset that was systematically analysed for the current study. In total, 38 students (registered Year 4 students in the respective academic year) were enrolled in the course, out of which 29 were female and 9 were male, and 16 were “locals” (i.e., UAE nationals), and the rest were “expat residents of the UAE” from the following countries: Egypt, India, Iran, Iraq, Italy, Jordan, Nigeria, Palestine, Sudan, and Syria (listed in alphabetical order). The enrolled students were aged either 22 or 23, with only one student, as an outlier, who was 26 years old. Out of the 38 students enrolled in the course, 37 submitted a reflective essay. The students were asked to give their written informed consent that their reflections will be used for research purposes. All the students who submitted a reflective essay approved to participate in the current research study (no.= 37). To protect the anonymity of the participants, all names were removed, and instead each submission was assigned a number serially (01 through 37, followed by “F” for when the participants are female and “M” for when they are male).

### Data Analysis

The data analysis began after the conclusion of the data collection phase. The data was inductively analysed, by two researchers (B.N. and F.O.), using a participant-focused, Phenomenological approach to thematic analysis [31]. The factors that could influence the researchers’ perceptions regarding the subject matter were recognized upfront. Consistency, throughout the analysis process, was assured. This iterative approach was based on the constructivist epistemology [32, 33]. In contrast to traditional scientific research, this interpretative understanding process required the recognition and recreation of the participants’ lived experiences. This approach is not concerned with finding casual explanations but rather to understand the participants, and their attitudes, behaviours, and actions [34]. This methodology assumes that we can capture what participants think by interpreting and in turn developing a thorough understanding of their self-expressions[35]

The process of analysis followed the six-step framework initially introduced by Braun and Clarke [36]. This multi-phased approach to inductive qualitative analysis is encouraged in socio-behavioural research, in general, and health professions’ education research, in specific [18, 37]. NVivo software version 12.0 plus (QSR International Pty. Ltd., Chadstone, Australia) was utilized to code the data, and in turn accelerate the classification of the identified text segments.

The first step of the analysis process related to the two researchers familiarizing themselves with the data. In the second step, the transcripts were reviewed while text fragments that relate, directly or indirectly, to the research question were extracted. This kept going on until no additional insight was generated from the datasets, and as such it was determined that data saturation was attained.

This led to the identification of categories of text fragments which set the stage for the researchers to initiate the third step of the analysis. As such, these categories underwent several rounds of reflections; all the potential ways by which codes of these categories could relate to one another were identified. Then, as part of the fourth stage, the researchers found the best way to merge the categories into higher order themes (Figure 2). All the themes and categories were then defined, in the context of the study, to complete stage five. The output of this step constituted the study’s conceptual framework which guided the last step of the multi-phased thematic analysis: reporting upon the findings, which was done narratively in alignment with established guidelines [38-40]. To further substantiate the findings, the researchers conducted a tally and reported on the number of text fragments within each category of the identified themes. If for a single participant, more than one relevant text fragment was identified, they were all collectively considered one entry. In other words, the tally is actually reflective of the numbers of participants that reflected on matters relevant to the respective categories.

**Figure 2.**
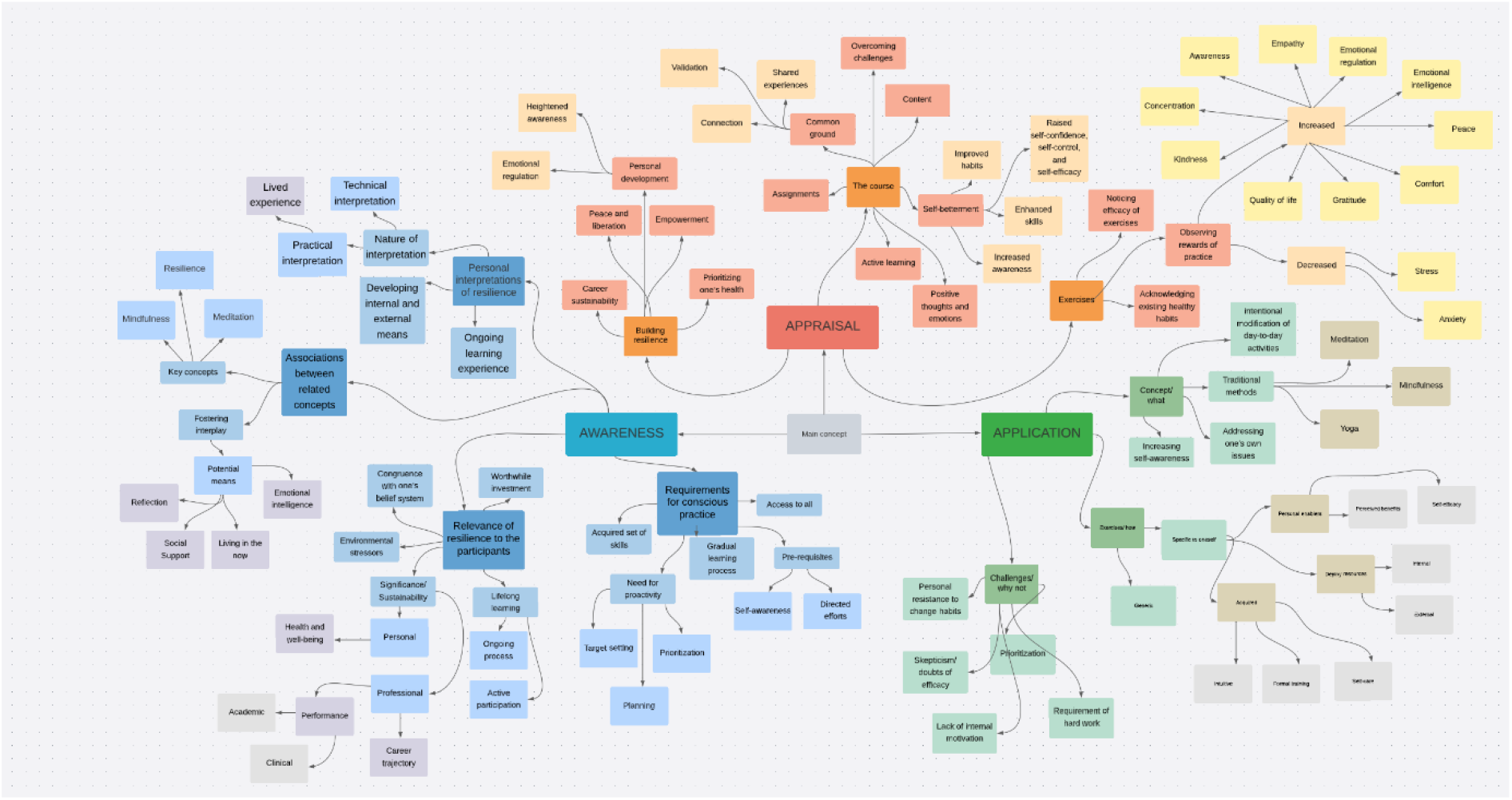
Mind map deployed as a tool to facilitate the qualitative analysis

**Figure 3.**
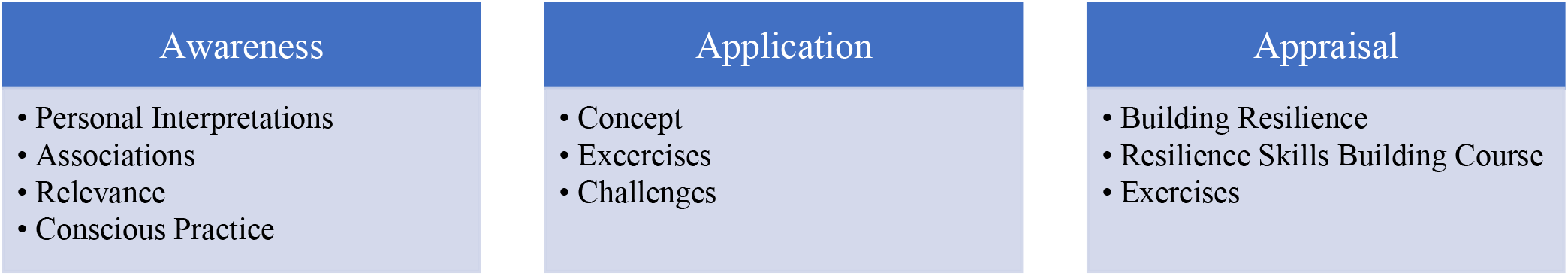
The study’s conceptual framework: 3As of Building Resilience

## Results

The qualitative analysis generated, as per this study’s conceptual framework: 3As of Building Resilience (Figure 1), three interlinked themes, namely: Awareness, Application, and Appraisal.

Within Awareness, four categories were identified: Personal Interpretations of Resilience, Associations between the Related Concepts, Relevance of Resilience to the Participants, and Requirement for Conscious Practice. As for Application, it included: Concept (What?), Exercises (How?), and Faced Challenges (Why Not?). Lastly, within the third theme: Appraisal, the following categories were identified: Building Resilience, Resilience Skills Building Course, and Exercises. The tally of text fragments showed the following distribution: Awareness (no.= 64), Application (no.= 46), and Appraisal (no.= 44) (Table 1).

**Table 1.** Semi-quantitative tally of the output of the participant-focused qualitative analysis

| Theme | Awareness |  |  |  | Application |  |  | Appraisal |  |  |
| --- | --- | --- | --- | --- | --- | --- | --- | --- | --- | --- |
| Category | Interpretations | Associations | Relevance | Practice | Concept | Exercises | Challenges | Resilience | Course | Exercises |
| Tally | 24 | 11 | 21 | 8 | 9 | 26 | 11 | 18 | 14 | 12 |
| Sum | **64** |  |  |  | **46** |  |  | **44** |  |  |

### Awareness

This theme encapsulates fragments of text that highlight the participants’ understanding/ cognizance of the concept of resilience, and its relevance to their personal and professional trajectories.

#### Personal interpretations of resilience

This category revolves around text excerpts that show the students’ individualized explanations of how one can develop the internal and external resources to handle life’s challenges.

> 6F: “…’toughness’ is not a single attribute of an individual, but rather a result of numerous internal qualities and external resources…”
>
> 15F: “…it allows people to remain level-headed during a crisis and to be able to move on from the incident minimizing negative long-term implications…”
>
> 20F: “…Resilience is the mental reservoir that grants us the psychological strength to push through the endless flow life stressors …”

Some students seem to interpret resilience in a broad sense.

> 1M: “…resilience to me refers to willpower. The ability to persevere when times get tough…resilience is what gives people the psychological strength to cope with stress and hardships…”
>
> 4F: “…I realized that it is not about ‘not experiencing hardships’… Life is hard. Resilience enables you to bounce back from the problem and not remain ‘stuck’…”
>
> 18F: “…resilience is actively cultivated to equip individuals with the conducive coping strategies to weather any adversity in a healthy manner …”

Other students seem more practical. Some of the interpretations appear to be based on lived experiences.

> 12F: “…conscious decision to keep going, despite all the challenges that one may face…resilient people have a compelling reason to get out of bed in the morning every day as they are committed to their life and goals…”
>
> 14F: “…resilience enables us to keep moving forward with intention and meaning rather than only for the sake of moving forward by helping us to integrate our experiences and thus re-evaluate our approaches every living moment…”
>
> 17F: “…it is knowing when to stop, acknowledging one’s limits, and remaining aware of when to take a break…”

Students appear to perceive building resilience as an ongoing learning experience.

> 13M: “…resilience can be seen as an active, continuous process, rather than a one-time adaptation or adjustment following a stressful experience…”
>
> 19F: “…resilience is not a skill that we are born with. It is a characteristic that one learns to develop and builds slowly over time; a set of skills that we continue on developing throughout our life…”
>
> 32F: “…resilience, for me, means the ability to adapt to life challenges. It is the plasticity and flexibility that allows me to overcome hard feelings and tough moments. Resilience is understanding the importance of self-care including that of our physical and mental health. It is the small steps that can be done every day and that eventually lead to ‘big’ outcomes…”

Perceived associations

This category highlights the reflections of the students in relation to resilience, and other contributing factors, such as: mindfulness and meditation, and the interplay across them.

> 18F: “…A variety of types of meditation exists, ranging from transcendental meditation, yogic meditation, progressive relaxation meditation, chakra meditation, and visualization meditation among many others-all sharing the same fundamental principle: to be able observe one’s thoughts from a neutral perspective…”
>
> 22F: “…I would like to think that resilience starts with mindfulness; to be resilient in difficult situations requires a moment of reflection, and awareness to your internal reaction and in turn external response to the obstacle…”

Mindfulness and meditation, along with their perceived interplay, are frequently alluded to when considering means of fostering one’s resilience. Some students highlight the importance of the practice of reflection, in relation to mindfulness and/ or meditation.

> 1M: “…meditation nourishes and expands mindfulness…”
>
> 6F: “…reflection is also important for mindfulness, it permits us to venture outside the circumstance, considering ourselves to be the onlooker as opposed to the person in question, and frees us up to different perspectives. By widening our point-of-view, we are better prepared to manage reactions that are usually ruminous…”
>
> 20F: “…to practice mindfulness, we can train ourselves to stop rehashing the past in our thoughts or imagining the future, and actually be in tune with what we sense presently…”

The sensory experience of ‘being present in the moment’ is repetitively mentioned in relation to mindfulness.

> 17F: “…mindfulness is the practice of remaining truly and wholly present in one’s reality…”

Social support is also pinpointed as a protective factor when it comes to handling challenges and fostering resilience.

> 1M: “…Mentally strong people tend to have the support of family and friends who help bolster them up in times of trouble…”

The students also appear to view emotional intelligence as a variable that plays a crucial role in building resilience.

> 15F: “…another variant that contributes to resilience is emotional intelligence which seems to be a very important concept when it comes to building resilience and practicing gratitude…”

#### Perceived relevance

This category encapsulates the text fragments that show in what ways building resilience is significant to students’ personalized experiences, be it in the professional or personal realm.

> 4F: “…It helps to slow-down our fast-paced lives and minds…”

In general, it was clear to the students that building resilience skills is in congruence with their belief that investing in preventative care has ample long-term, protective benefits.

> 13M: “…as medical professionals, we believe ‘prevention is better than cure’, which is why building resilience makes sense to us… as students, we must focus on becoming resilient by developing the emotional and mental skills necessary to prevent outside pressures from hindering our perceived potential, even before we are faced with stressors…”

The students seem to comprehend that resilience is not a static status that one attains, but rather an ongoing process that requires active participation, which they are directly associating with continuous growth and lifelong learning. The perceived meaning of resilience appears to evolve as the students are progressing in their educational journey.

> 2F: “…the meaning of resilience to me changes with each stage of my medical career…”
>
> 20F: “…In every stressful situation, by finding and focusing on the opportunities for growth, we enable ourselves to remain calm and constructive…”

The necessity of building their internal reserves is frequently alluded to be it in relation to their education, and/ or their future as professionals in the medical field.

> 5F: “…as future physicians, we will inevitably come across many challenges on a regular basis. It may be easier to shy away from these challenges, but with resilience, we are able to face the challenges head-on, and better even endure the situation…”

The benefits of building resilience in relation to their health and wellbeing is brought-up, where the students reflect on its potential in reducing stress and anxiety.

> 9F: “…as medical students, we go through a lot of stress and anxiety; mindfulness is one of the most efficient ways to reduce stress and go about our days in a relatively good mood… I think mindfulness is a vital technique that helps with focusing… accepting and appreciating our surroundings…”
>
> 19F: “…practicing mindfulness and enhancing resilience are beneficial tools that help mitigate stressors and burnout which improve the wellness of medical students…”

Building resilience seems to be particularly relevant to the students’ learning experience. The participants highlighted the aspects of academia that are burdening them including but not limited to study load and transitions, all of which they believe affects their performance. Preventing academic burnout is clearly pinpointed in the students’ reflections.

> 1M: “…this is particularly pertinent when studying medicine as the study load can at times become quite burdensome…”
>
> 10M: “…as a medical student, it is no surprise that resilience is quite important in order to be able to cope with the stress and anxiety of academia which can be overwhelming… practicing mindfulness and building resilience skills are imperative to avoid burn-out and to maintain one’s wellbeing…”

The students are aware of the benefits of resilience in relation to the transitions that are integral to their career trajectory.

> 12F: “…Transiting from the classroom to the clinical environment holds a lot of new challenges and can trigger many mental health conditions, and practicing resilience could play a protective role in clinical training and can help improve our professional quality-of-life during our clinical rotations….”

It was obvious to the students that resilience contributes to paving a sustainable pathway, since as future physicians the students believe that they will inevitably encounter challenging, unpredictable situations, pertaining to patients’ lives.

> 12F: “…the medical profession also involves inherent unpredictability that demands future doctors to be adaptable, therefore, resilience is one of the most valuable traits a physician can have to enhance psychological strength to cope with stressful situations in our life…”
>
> 18F: “…life as a professional in the healthcare sector can be remarkably arduous and demanding, physically and psychologically. Enduring long working hours, day-to-day exposure to patient traumas and deaths, the looming overhead fear of contracting infectious diseases are just few of the numerous stressors that tie into how truly unpredictable a regular workday can be for a physician…”

It also contributes, according to the participating students, to the physicians’ clinical performance and ultimately the outcomes of care.

> 19F: “…building resilience among doctors and healthcare workers has a long-term effect on the quality of health care, and strongly impacts the way in which patients are cared for…”

The relevance of resilience is also brought-up in relation to the attributes of the external environment, such as: today’s fast-paced life, the more recent COVID-19, and the ongoing pressures of the modern world.

> 18F: “…these stressors are more than sufficient to induce detrimental psychological reactions, and result in burnout… resilience has never been more relevant among healthcare professionals, given the current, unprecedented pandemic…’

#### Perceived requirement for conscious practice

This category sheds light on the students’ awareness that resilience needs proactive building through directed efforts.

> 24M: “…I hope to master mindfulness to proactively use it to face everyday struggles that occupy my mental space. At the end, it is not the destination that determines success, but the small victories throughout the journey…”

It is clear to the students that resilience is not innate.

> 12F: “…resilience is not a trait that we are born with; it is a set of skills that one learns to develop and build slowly over time…”

In fact, the students appear aware that building the associated set of skills requires proactive measures such as: setting targets, planning, time management, and prioritization.

> 4F: “…I tried this for two weeks; I have set a plan to build my resilience by practicing mindfulness, prioritizing my relationships, and keeping things in perspective…”
>
> 7F: “…in a moment of stress, I thought about trying out a mindfulness activity, there in the moment…”
>
> 36M: “…praying on time, playing football, going out with my friends, and spending more time with my family, without affecting my studies, were the initial changes in my lifestyle… I prepared mindfulness objectives, for each week, and tried achieving them day by day…”

The students repetitively mention that resilience is not unique to any one person. To them, the skills of resilience can be acquired by anyone provided there is self-awareness, compounded by an investment of efforts in the right direction.

> 16F: “…if you can identify the stressors in your life and not just sit idle but really try to untangle this mess, you would actually feel lighter…”

It is apparent to the students that resilience is not acquired suddenly as a one-time thing. It rather requires a process of learning, where the set of skills are gradually, with intention, built over time.

> 1M: “…people are capable of learning the skills to become more resilient…”
>
> 2F: “…I do believe that once a person knows what resilience is and how to practice it, the act of building it over time would take a few seconds to minutes of a person’s time, and it would allow them to feel more at peace…”
>
> 35F: “…I started consciously bringing my awareness to any day-to-day task even during assisting in surgeries as part of my surgery rotation, and talking to patients in my psychiatry rotation…”

### Application

This theme refers to the participants’ personal reflections on the practical aspects of resilience.

#### Concept (What?)

This category shows the students’ understanding of the differing means of building personal resilience. There is frequent mentioning of the traditional methods (e.g., meditation, mindfulness, and Yoga), as well as reflections on the intentional modifications of day-to-day activities to infuse mindfulness into their lives.

> 14F: “…it is almost magical how versatile mindfulness is, taking innumerable shapes and forms, and influencing our lives in diverse ways. It can be incorporated into anyone’s lifestyle, through particular activities, including ones that address physiological cravings and consequential behaviours…”
>
> 2F: “…a person can build resilience through meditation and mindfulness…”

In some instances, the students refer to the commonly used labels of the traditional methods, and in other situations, they choose to further elaborate on how those methods exhibit themselves in the context of their personal lives.

> 5F: “…Mindfulness allows one to get in-sync with their breath. This helps to increase one’s awareness and enhances their state-of-mind…”
>
> 22F: “…being resilient and mindful is crucial to have a healthy ‘academics-life balance’… acknowledging that difficulties in life are inevitable and remaining mindful that everyone around us is facing some sort of difficulty help me to somehow find a way tackle whatever situation I find myself in…”

Self-awareness is emphasized as one of the means of building resilience. The students appear to associate self-awareness with personal and professional development.

> 3F: “…the practice of mindfulness helped me become more self-aware by observing the flow of inner thoughts and emotions. I now recognize my emotions and their triggers, and instead of burying my head in the sand as was my previous custom, I am now able to face my anxiety head-on…”
>
> 13M: “…Mindfulness also allows us to be aware of what is going on inside and around us, providing a clear picture of our situation…”

Students clearly communicate that addressing their own internal issues have contributed to building their own resilience.

> 28M: “…with respect to mindfulness, I have been able to understand a lot of insecurities and issues I had, which were leading to my troubled mind; it has also helped me understand how to deal with people and with situations I feel uncomfortable with…”
>
> 5F: “…as a person who has always doubted oneself and was my own worst enemy, I built resilience through silencing my internal voices of self-doubt… I built myself back up and started afresh…”

#### Exercise(s)/ Activities (How?)

This category puts emphasis on how the students are actually putting into practice what they have identified as means to build resilience.

> 12F: “…simple acts of meditation and mindfulness like the five senses exercise can help us to adapt to the new environment we are in and to the new stressors we are facing to orient to our environment instead of focusing on our racing thoughts…”

Among the identified exemplars are ones that indirectly show the personal factors that enable change in behaviour. These include what the students perceive as the (short- and long-term) benefits of changing their behaviours and how efficacious they believe themselves to be.

> 20F: “…Traditional methods of contemplation can also be tools such as yoga and meditation but developing the awareness to one’s breath can truly be a game changer…”

The students reflect on instances where they deploy their internal resources and others where they rely on external ones.

> 10M: “…sometimes additional support from a friend, family member, or counsellor may also help in building resilience…”

Some of the activities, that the students reflect upon, were adapted through formal training, and others were naturally acquired or intuitively performed.

> 2F: “…this can be done through simple activities, such as the five senses exercise, which could calm a person down during periods of anxiety…another way to practice mindfulness is through building awareness of one’s own breathing…”
>
> 4F: “…during one of the course sessions, we were led to eat a date mindfully; it was as if we are eating a date for the first time ever…”

In some instances, the students reflect on activities that are specific to them.

> 2F: “…I choose to walk around, either at home or outdoors, allowing myself to let go of life’s stressors…”
>
> 3F: “…my first experience with mindfulness was simply by observing my backyard garden…”
>
> 25F: “…I started implementing extra-curricular activities like jogging and dancing to make myself more comfortable and less stressed…”

In other instances, the students provide generic explanations including definitions and/ or scientific explanations of how the particular technique affects one’s health and wellbeing.

> 9F: “…practice of reflecting is also very crucial, it helps one to step beyond the situation and exercise viewing it differently, it gives you the ability to be a spectator…”
>
> 10M: “…some of the mindfulness exercises include focusing and paying attention to what is at hand, focusing on the present moment, accepting oneself, and focusing on breathing, meditation, yoga, exercising…”

Most of the students coin the term mindfulness, associating it with various activities that promote living in the moment, and adopting acceptance and a non-judgmental attitude.

> 15F: “…I have been using the method of breath meditation for a few years, on my apple watch, and I can attest to the fact that it provides an immediate sense of relaxation…”

The students repetitively allude to instances where they reframe their cognitions.

> 8M: “…one thing that I always do when I feel down is looking back at myself a few years ago and asking, ‘aren’t there many things that you wished for back then that you now have?’ This question never fails to make me feel better…”
>
> 10M: “…I have been effectively reframing my thoughts by not viewing a glass as ‘half empty’ but rather as ‘half full’, and focusing on what is within our locus of control…”
>
> 14F: “…I have, over the years, intuitively learned to slow down and re-consider the situation with fresh perspective to truly master any difficulty, and looking back at this now, I see mindfulness in this practice, even without having had any formal education about it…”

The students also mention activities which have a predominant physical element and solicit change in bodily sensations.

> 29F: “…one way I personally find useful is removing myself from the situation or stressor to be able to calm down, basically unplugging myself from the source of fuel. Taking a walk to reorganize my thoughts is something I find particularly beneficial or going to the gym to let off some of that built up steam…”
>
> 30M: “…apart from a lot of the breathing exercises and relaxation techniques, one method that has greatly helped me has been body scan meditation…”

It appears to be clear to the students that they need to take good care of themselves to maintain their mental health.

> 10M: “…I personally find several things that help me such as: relaxing when I feel tired; knowing my limits, and not pushing myself beyond them to avoid burning out; effectively managing my time; listening to music; and sometimes just sitting in silence and peace for a few minutes…”
>
> 18F: “…recharging before facing a new challenge, taking a break from routines, and using humour in stressful situations… religious or spiritual means that help build resilience…”
>
> 33F: “…it is important to me that I have adequate sleep and eat a variety of nourishing foods to feed the mind, body, and soul…”

The students are also aware that building resilience is a process, and the proficiency of any one technique (e.g., mindfulness) and/ or changing subconscious patterns require time, intention, and perseverance.

> 15F: “…after becoming more familiar with the concept of resilience, I moved to practicing it in my daily life…”
>
> 18F: “…there are many strategies designed to help nurture mindfulness, such as, paying attention to each of the five sensory experiences…”
>
> 26F: “…I have been practicing how to effectively use my time by learning to strike a balance between studies and leisure… Upon reflection, I have realized that I used to engage in various maladaptive coping mechanisms such as avoidance and denial… Attempting to become more aware of my choices when dealing with difficult situations has enabled me to come out the other end more resilient than before…”

#### Faced challenges (Why not?)

This category pinpoints the statements that the students used to reveal what they perceive as the direct or indirect obstacles to engaging in resilience skills building exercises. In some cases, the students shed light on their own resistance towards changing old habits and adapting new ones.

> 6F: “…practicing mindfulness is something that probably a lot of people have on their daily to-do lists, but never get around to it because, they either do not believe it works or they are too busy with their lives to stop for literally just a minute to breathe…”
>
> 31F: “…It is all easier said than done…”

Some students implicitly state that mindfulness exercises seem to them to be too simplistic of a technique to be able to help them build resilience. In other words, they had doubt about the efficacy of the techniques. In some cases, scepticism was expressed.

> 9F: “…practicing mindfulness can be considered, by some, as ‘difficult’ and ‘a waste of time’…”
>
> 27F: “…To be frank, upon first encountering mindfulness, I was nothing short of sceptical: ‘how could the solution to something so troubling be simply to stare or contemplate?’ …”

Contradictorily, some students seem aware of its efficacy but the challenge for them was more around making it a priority in their lives. Moreover, the students express how making active changes discouraged them which could signify lack of internal motivation.

> 11F: “…I do not regularly practice meditation, because firstly, I do not have time or rather do not remember that I have the option to sit and meditate when I get the time, and secondly, I do not feel the need to practice meditation…”
>
> 35F: “…resilience and mindfulness were things that I never really had the time for…”

There is also a perception among a few students that building resilience skills requires a substantial amount of effort. This seems to contribute to the students’ limited engagement in the suggested activity, despite their knowledge of its efficacy.

> 20F: “…however, learning how to pay attention to my thoughts and tune into my emotions without having clouded judgment towards myself proved to be quite a tedious task…”
>
> 22F: “…I have had many days in which I needed to be resilient but found it easier to just succumb to the anxiety and not do anything about it…”
>
> 28M: “…initially, when one starts with exercising mindfulness, it can be really tiring…”

### Appraisal

This theme represents the participants’ evaluation of resilience, and of the corresponding skills, whether acquired through the respective course (or another formal training), or otherwise.

#### Building resilience

This category highlights the students’ reflections on the value (or its absence) of proactively developing one’s mental health.

> 21F: “…I started meditating every day for a full month. My personality started to change for the better, I became less stressed… the main advantage that I gained from resilience was how to manage my health… it has improved my physical and also my mental health…”
>
> 34F: “…The importance of resilience and mindfulness cannot be overlooked, especially among medical professionals… It is essential for everyone to be equipped with reliable coping mechanisms if they wish to maintain a good mental health…”

There appears to be, among the students, recognition of the value of developing resilience as a form of consciously helping oneself cope with life’s challenges.

> 14F: “…only recently have I become proactive about my wellbeing and interpersonal relationships, thanks to mindfulness…”
>
> 28M: “…thanks to mindfulness, it has become easier for me to take criticism and openly share my opinion…”

The students seem to acknowledge that with resilience comes heightened awareness and emotional regulation, as well as empowerment that allows one to feel more at peace and liberated.

> 3F: “…to utilize my own consciousness to battle the many emotional challenges that I tend to experience, is a healthy coping mechanism and an empowering way to declare independence. For that, I am grateful…”
>
> 7F: “…I think I want to make mindfulness a habit… Mindfulness helps me live in the moment and this makes me feel better…”
>
> 16F: “…I discovered that I can calmly analyse and figure out what is bothering me, and why I feel the way I feel… it gave me a superpower, which I use for my own good…”

Students mention becoming more conscious of their health, making it a priority, and this they attribute to their increased awareness that (in their opinion) is a direct result of proactively practising mindfulness.

> 6F: “…practicing mindfulness, especially during challenging times, allows me to be calmer and more peaceful, and it makes me more aware of my emotions so that I can better regulate them…”
>
> 21F: “…I was able to cope with stress healthily, so stressful situations stopped affecting my mental and physical health like they used to do…”

The students also refer to the connection between building resilience, and continuity and sustainability in their chosen field of study, namely: medicine.

> 25F: “…If I do not practice mindfulness, I think I would have burnt-out early on in medical school, and I would have had a very tough time navigating my medical school commitments, along with my personal and social life…”
>
> 32F: “…I have become more flexible to withstand all the pressure that I am encountering in the hospital… I am not scared of making mistakes anymore; this has enabled me to try new things and learn more each day… it enabled me to understand that anything that helps my mental health is just as (if not more) significant as my studies…”

#### Course

This category shows the students’ perception of the structured learning experience, within the curriculum, which they were required to complete as part of the respective MBBS (i.e., the intervention under investigation).

> 15F: “…this course aided us in acquiring a skill that I believe is essential to life…”

It was evident, from the exemplars, that most of the students highly value actively engaging in the course and with its content. Students appreciate the assignments requested as part of the course, which they believe, has been instrumental to having them make changes to their habits and lifestyle. Some of the students even mention that a few of those exercises eventually became second nature.

> 6F: “…having this as an actual assignment was exactly what I needed to get a jumpstart on mindfulness techniques in my daily life…”
>
> 26F: “…the resilience course made me far more aware of the way I have been dealing with life, and its stressors, all these years… the weekly mindfulness sessions have been able to point me in the right direction when coping with my daily struggles… I have been focusing on the present moment rather than worrying about what has happened or is yet to happen. I achieved this by combining various techniques such as meditation and muscle relaxation…”

It appears that bringing the students together for this course somehow create a common ground among them, where they feel connected and validated through their shared experiences.

> 4F: “…I also realized that I was not the only one going through a rough time mentally, which made me feel less ‘alone’. Also, at the end of each session, I always felt better and relaxed…”

Several students emphasize the emotion of gratitude in their personal expressions (either in relation to experiencing the course and/ or as a habit acquired through exercises practiced as part of the course).

> 25F: “…this course was delightful; it made my colleagues and I more open to constructive thinking… it encouraged me to figure-out solutions to my everyday problems and struggles…. it inspired me to be more thankful for the good things around me and less judgmental of the things I think are bad…”
>
> 26F: “…another key aspect that caught my interest during the mindfulness sessions was the concept of gratitude… one of the course activities required that I write-down in separate pieces of paper the things that I am grateful for and to place these papers in a small jar. When I experience difficult times or feel low, reading one or more of what I had identified somehow lifts me up…”

Appreciation is frequently alluded to in relation to the skills acquired via this learning opportunity.

> 11F: “…One thing I learnt from this course is to accept my negative emotions, like anger, anxiety, and stress, and acknowledge them…”
>
> 14F: “…I truly believe in the importance of resilience, and it is only through this course that I have been exposed to the different methods of strengthening this prime skill…”

It seems that the evidence-based course content contributes to enhancing the students’ understanding of mindfulness and in turn lessen their scepticism about it. The students corroborate on the efficacy of the skills learnt by citing examples from their daily lives of how their thoughts, behaviours, and feelings have been positively influenced.

> 4F: “…when starting the resilience classes, I realized how easy it is to do and that it is not time-consuming… All the points covered in the sessions were backed up by research, which made me eager to start and change the way I live my life daily….”
>
> 7F: “…the resilience course made me learn that meditation is not about not getting distracted at all, but it is about bringing yourself back to the moment when you get distracted…”

Students appreciate that the course equipped them with a set of lifelong skills which they believed will help them to sustain their career in the long run. It is frequently mentioned that the course has been a catalyst for an increase in awareness, and acknowledgment of habitual behaviours that are self-sabotaging. There are instances where students credit the course to an increase in their self-confidence, self-control, and self-efficacy.

> 26F: “…This technique has also made me aware of my tendencies to focus on the negative, to judge, and to generalize… Overall, this course has developed my capacity to maintain my own mental health, which I am hoping will make me a better healthcare professional…”
>
> 32F: “…during the first few weeks of my rotation, I was always worried about how I am performing. I had little confidence in my abilities and was scared most of the time that I am not meeting the expectations of the supervising doctors. During that time, every little mistake I did would put me in a very bad mood for few days and would make me doubt everything I know. Consequentially, I was not able to learn from my mistakes; they represented bad experiences rather than opportunities to identify my weak points and work on them. This mindset has changed drastically because of the resilience course…”

Apparently, routine practices of mindfulness and skills learnt during the course have been incorporated into daily life, by some students, not only on a personal level but professionally, as well, during clinical rotations.

> 35F: “…this course gifted me new coping skills that I can deploy at any time to handle the challenges that I face, and in turn, come-out stronger… this course offered me a new perspective, problem-solving skills, made me more aware of my surroundings, and helped me let go of many of what constituted barriers between me and my goals. It gave me the strength to wake-up every day, and attend to my duties and responsibilities not just as a student, but as a friend, daughter, sister, and a human…”

#### Exercise(s)

This category uncovers the students’ impressions about the exercises that they have pinpointed in terms of building one’s resilience (whether acquired through the respective course or otherwise).

> 6F: “…it is such an easy task, and it takes less than 5 minutes. It can turn around your entire day…”

Most of the students reflect upon the influences of activities that they realize one needs to be proactive about developing.

> 18F: “…meditation can be seen akin to bathing the physical body; it cleanses the mind… it purifies the mind, allowing one to start each day afresh, with a clean slate…”
>
> 34F: “…I try to get some yoga into my schedule. It reminds me to listen to my body and take care of it… all helps me stay relaxed and mindful of my blessings…”
>
> 36M: “…. I became mindful of the details… My sense of contentment significantly increased…”

Also, few of the students reflect upon how they became more aware of certain healthy habits that they had subconsciously adapted over time to cope with the challenges in their lives.

> 23M: “…I have opened myself to many new experiences that allows me to become one with the moment, such as enjoying a walk and doing some Tai Chi… I limited mindless scrolling through my social media accounts, and procrastination…”
>
> 25F: “…I practice mindfulness while working-out. Running has always been a great stress-reliever for me: I imagine that I am running away from negative thoughts and towards better ones. I also enjoy the sense of accomplishment I experience after a good run …”

A lot of the students emphasized how those exercises help them ‘go back to themselves’.

> 11F: “…I try to spend 5 minutes at the end of each day before I sleep to count my blessings and thank God for all of them…”

There is a prominent acknowledgment of the efficacy of the mindfulness exercises (in particular, the ones that include sensory organs and the whole body) since an immediate shift in energy tends to be experienced.

> 20F: “…I have personally found that mindful breathing and prayer help achieve inner peace… I have come to learn that practicing mindfulness involve acceptance and a non-judgmental mindset…”

The rewards of regular mindfulness practise at home have been linked, by the students, to improved sleep and concentration, and to a decrease in stress and anxiety levels.

> 37M: “…I did that for about 2 weeks, and my sleep schedule got adjusted and worrying about the future lessened. Meditation is a helpful skill that works for me when I need it…”

Some students highlight how the practise of the exercises help them better regulate their emotions which has contributed to heightening of their emotional intelligence, thereby increasing their empathy and kindness towards other people.

> 30M: “…I do believe that I have become a kinder, calmer, and more patient person since I started resorting to mindfulness and meditation to manage my stress…”
>
> 34F: “…has helped me increase my ability to regulate my emotions as well as decrease the amount of daily stress I deal with. In addition to affecting how I view myself and my life, mindfulness has also improved my emotional intelligence and my ability to relate to others with kindness, acceptance, and compassion…”

These exercises, including but not limited to: praying, are considered by the students as anchors during troubled times, helping the students attain inner peace, comfort, and increased gratitude.

> 3F: “…As familiar as I presumed myself to be with the scenery, I found myself noticing the simplest yet most intriguing details. For once, I dwelled upon the trees and greenery instead of the scorching hot sun of Dubai. It was astounding how focusing on such robust objects produced an immense amount of relief.

Instantaneously, I could sense physiological changes within my own being and a shift of consciousness…”

> 4F: “…practicing mindfulness in my life has brought me a lot of comfort as it helped with my anxiety and my overthinking tendencies…”
>
> 20F: “…for me, leaning on faith and having that spiritual mission mean having peace of mind, stability, and the security of knowing that I have my own compass to guide me when the going gets tough…”

Students mention a slowdown in their fast-paced lives. This, they suggest, has opened them up to new experiences and has allowed them to enjoy even fleeting moments that they had been previously taking for granted.

> 7F: “…It also helps me to be grateful for even the ‘little’ things around me and in my life. I also find that I feel more balanced and can better respond to situations around me…”
>
> 25F: “…my workdays are usually fast paced yet, practicing mindfulness slows me down…this helps me clear my mind; I find it very relaxing… I enjoy practicing mindfulness in the morning right after waking up. I soak in all the sunshine from my bedroom window…”

## Discussion

The current study shows that effectively building resilience skills is appreciated among medical students. They believe a curriculum-based resilience skills building course offers them immediate benefits as well as long-term impact. Such an intervention improves the health and wellbeing of the students, as per their reflections. In addition, the students perceive their capacity to develop meaningful relationships to increase as a result to building resilience skills. They also think that it improves their academic performance and maximizes their learning. According to the students, and in congruence with the literature around the subject matter [18], actively co-creating their learning experiences, as part of such an intervention, adds to their confidence, making them self-regulated learners which is, at the present time, expected of students who are at university level.

One’s self-efficacy is instrumental in determining whether coping behaviour will be initiated, the amount of effort that will be utilized, and how sustainable the behaviour will be during challenging experiences [41]. A triadic interaction between behaviour, and personal and environmental factors facilitates the entailed learning, popularly known as reciprocal determinism (Bandura’s model). The resilience skills building course, investigated in this study, is reflective of this concept, where it was designed to raise individuals’ confidence and their self-efficacy. It also aims for behaviour change by offering assistance and social support in the form of group learning but with the cognizance that environmental and personal constraints could be deterrents to behavioural change.

According to the participants of the current study, building resilience plays a significant role in nurturing competent, future-ready physicians, which in turn enhances the quality of care and patient outcomes. The dimensions of resilience include, but are not limited to: self-efficacy, self-control, help seeking behaviours, learning from adversities, and perseverance despite the challenges [42]. Building those skills is considered good practice in contemporary medical education. However, officially integrating the development of these competences, as part of the medical education curricula, is yet to be implemented among medical schools in the MENA, in general, and the UAE, in specific.

Since the COVID-19 pandemic, health care as an industry has been undergoing a significant paradigmatic shift to focus more on proactive pre-emptive care for well-being [43]. This demands a new meta-competency to make transformative changes in times of exceptional complexity and ambiguity [44]. By incorporating the resilience building course into the curriculum, anchoring its design in constructivism experiential learning theory which views learning as a process of active adaption [27, 28], the students had the chance to engage in reality-based problem-solving contexts.

A major characteristic of the resilience course was the structured reflection time, at the end of every session, which is an integral attribute that is expected from all university students [45], even more so with medical students, particularly during their clinical placements. The ‘soft’ skill of reflecting is considered to be the cognitive bridge between what is learnt in the classroom and its practical application in the real world [18], empowering them to trust their personal insights. This enhances their emotional flexibility as well as their ability to critically analyse complex issues which is essential in their field of work for the larger good of patients in the future. In fact, most of the participants of the current study seemed idealistic in their responses and very reflective. The content of their write-ups appeared quite sophisticated.

The substantial capacity to reflect that was evident in the content of the essays can be attributed to the students’ acknowledgement of the importance of reflecting on experiences as an exercise which builds one’s resilience, as pointed out by the students themselves. Also, it is worth noting that the students’ perspective of mental health seemed quite holistic, and not limited to cognitions, emotions, and behaviours. They repetitively refer, in their narrations, to the body, bodily sensations, and shifts in energy, which is linked to the extensive works around increasing one’s sense of awareness of internal experiences [e.g., Somatic Experiencing® (SE) [46]].

Through this course, mindfulness was utilized to scaffold reflection. With a growing presence of groups of mindful, eager, and critical intellectuals who are choosing to enhance their executive functions of autonomy, emotion regulation, and better relationship capacities [47], the course is an attempt to help them make informed decisions and actions, particularly during challenging conditions. It justifies why students have frequently alluded to mindfulness, in their reflective writings, as a “foundation skill”. The explanatory nature of the generated data may be associated with the fact that the current study’s participants are medical students, most of whom are high functioning individuals. They appeared to praise activities that enable them to ‘get out of their heads’. Therefore, it is not unexpected for them to resort to definitions and scientific justifications.

The students have indicated an assortment of activities and habits that they proactively engage in to raise their resilience. These include ones that promote living in the now, physical activities and ones that solicit bodily sensation s, external resources that support in calming the nervous system, and ones which involve reframing of cognitions. These observations are in alignment with findings of a previously conducted systematic literature review that highlighted a combination of four intervention to be the most effective in promotion university students’ mental health, namely: Mindfulness, Movement, Meaning, and Moderator [15].

Students have also reflected on how the course fostered a sense of togetherness and collaborative learning which is grounded in constructivism theory [27, 28] that emphasizes participation in the social world as key to knowledge [48] highlighting the importance of contextual experiential learning [24]. The resilience course was uniquely designed in such a manner that students could actively engage in their own healing journey in a self-directed way to ensure the sustainability of the process making it a life-long learning. It is an attempt to guide students to move from peripheral participation to a more immersed one [24] by bringing-up examples of their own clinical experiences (in the sessions) which they have described in several instances in their reflective writings.

Students have highlighted the need to invest efforts as a major challenge which holds them back when exercising resilience. The required efforts, however, are mainly internal. For example, when one indicates: ‘we are used to ‘doing’ as opposed to ‘being’. This might be especially true for medical students as their demanding work-life might reinforce the habit of ‘doing’ over ‘being’[49]. Mindfully performing daily activities as part of their busy schedules involves learning to increase awareness without actively engaging with the content of the thoughts which they have mentioned as a challenge and a discomfort [50]. Habitual practises of having the mind wandering with the thoughts, indulging in attached observation [51], the physical discomfort of indulging in mindful practises, and self-doubt [52] have all been indicated as constraints to the practice. Practical limitations of feeling overwhelmed with grasping the new core concepts of mindfulness and prioritizing time for practice have also been mentioned by students.

To combat a few of these challenges, the delivery of the resilience skills building course was performed in small groups by dividing the class into two sections to enhance a sense of camaraderie and connection. This configuration was meant to foster the strengths of the participants, since in principle they belong to “Gen Z”, which a cohort known to be highly collaborative, self-reliant, and pragmatic [53]. The students were exposed to ‘McMindfulness’[54] wherein a reductionist element of the underlying foundational principle of traditional Mindfulness based Interventions (MBIs) was focused on. This was consciously incorporated, taking into consideration, the workload and time constraints of medical students which are often reported as deterrents towards longitudinal themes when compared to other disciplines [55].

The compelling need to promote medical students’ wellness is underscored by the concerning rise of emotional distress among them. There is enough evidence of the success of tools such as mindfulness meditation in enabling students to self-regulate to cope with stress and improve their psychological well-being [22, 56, 57]. The potential of mindfulness-based activities to rewire how the working memory and executive functions of the brain respond and revaluate stressful situations [58] is a skill that can be of immense benefit to medical students who by default work in dynamic conditions that require emotional and cognitive flexibility.

Despite the valuable evidence of the effectiveness of such resilience skills building courses, the paucity of well-designed curricula exclusively for medical students is still a challenge [59]. This is due to the practical curricular issues in terms of time restraint for this group of students due to demanding academic pressures that make participation in longitudinal programmes extremely challenging. There is enough scope to even integrate it into the mandatory formal courses as a pan university effort to encourage students to become more self-aware and to engage in self-care activities. This does not only equip them with skills to prevent burnout and compassion fatigue [60] but also enable them to become well equipped to foster wellbeing in the future among their patients. As such, they would become role models and walk the talk themselves.

One of the objectives of the course was to introduce resilience as a life skill that can help medical students to ground themselves to evolve into healthcare leaders who have healthy boundaries. This in turn will enable them to have a humble realistic estimation of their abilities and an acknowledgement of their limitations which will benefit not only themselves but also their future patients.

This study is characterized by a few limitations. Focusing on a single cohort from one medical university enabled the in-depth development of many thorough insights. Yet, the findings are only transferable to institutions which are contextually similar to that investigated in the current study. It would be worthwhile for future studies to investigate the perception of learners of similar interventions (aimed at building resilience) across several institutions. Moreover, the qualitative narrative data, compounded with the phenomenological participant-focused approach in the current study, allowed the researchers to tap into the students’ lived experiences which added significant value, in terms of the research findings. However, in terms of reliability of the methodology it would be worthwhile for upcoming studies to deploy a mixed methods research design that systematically merges the qualitative with the quantitative findings. Also, the current study helped in developing an impression of the efficacy of the intervention but not really its effectiveness. There is also the social desirability bias that might have affected the validity of the research findings, given that the students might have responded in ways that feel more appropriate. It would be interesting for future studies to assess the mental health status, using validated tools, of the students before and after the intervention to deem the effectiveness of the course.

## Conclusion

This study showed that integrating a resilience skills building course into medical curricula is likely to be positively appraised by the students, where it raises their level of awareness and likelihood of proactively applying the learned concepts in their daily lives. This is especially true when the course is anchored in constructivism experiential learning theory and designed to foster self-directed learning. This study provides support for further developing and implementing contextualized learning opportunities for building resilience skills among future healthcare workers.

## Data Availability

The data underlying the results presented in the study are available in the Supporting Information of the current submission.

## Notes

### Competing Interest Statement

The authors have declared no competing interest.

### Funding Statement

The author(s) received no specific funding for this work.

### Author Declarations

Ethical approval for the study was granted by the MBRU, Institutional Review Board (Reference # MBRU-IRB-2019-021). Written informed consent was obtained from all participants.

## References

1. Moffat KJ, McConnachie A, Ross S, Morrison JM. First year medical student stress and coping in a problem-based learning medical curriculum. Med Educ. 2004;38(5):482–91.

2. Rosenthal JM, Okie S. White coat, mood indigo--depression in medical school. N Engl J Med. 2005;353(11):1085–8.

3. Noreen A, Iqbal N, Hassan B, Ali SA. Relationship between psychological distress, quality of life and resilience among medical and non-medical students. J Pak Med Assoc. 2021;71(9):2181–5.

4. Anisman H. Stress and your health: From vulnerability to resilience. 2015.

5. Dahlin M, Joneborg N, Runeson B. Stress and depression among medical students: a cross-sectional study. Med Educ. 2005;39(6):594–604.

6. Dyrbye LN, Thomas MR, Shanafelt TD. Systematic review of depression, anxiety, and other indicators of psychological distress among U.S. and Canadian medical students. Acad Med. 2006;81(4):354–73.

7. Lin T, Stone SJ, Anderson T. Treating from Afar: Mental Health Providers’ Challenges and Concerns During the COVID-19 Pandemic. Behav Med. 2021:1–4.

8. Tang W, Hu T, Hu B, Jin C, Wang G, Xie C, et al. Prevalence and correlates of PTSD and depressive symptoms one month after the outbreak of the COVID-19 epidemic in a sample of home-quarantined Chinese university students. J Affect Disord. 2020;274:1–7.

9. Wang ZH, Yang HL, Yang YQ, Liu D, Li ZH, Zhang XR, et al. Prevalence of anxiety and depression symptom, and the demands for psychological knowledge and interventions in college students during COVID-19 epidemic: A large cross-sectional study. J Affect Disord. 2020;275:188–93.

10. Huckins JF, daSilva AW, Wang W, Hedlund E, Rogers C, Nepal SK, et al. Mental Health and Behavior of College Students During the Early Phases of the COVID-19 Pandemic: Longitudinal Smartphone and Ecological Momentary Assessment Study. J Med Internet Res. 2020;22(6):e20185.

11. Dyrbye LN, Massie FS, Jr., Eacker A, Harper W, Power D, Durning SJ, et al. Relationship between burnout and professional conduct and attitudes among US medical students. JAMA. 2010;304(11):1173–80.

12. Aburn G, Gott M, Hoare K. What is resilience? An Integrative Review of the empirical literature. J Adv Nurs. 2016;72(5):980–1000.

13. Jeste DV, Palmer BW. A call for a new positive psychiatry of ageing. Br J Psychiatry. 2013;202:81–3.

14. Folkman S, Moskowitz JT. Coping: pitfalls and promise. Annu Rev Psychol. 2004;55:745–74.

15. Nair B, Otaki F. Promoting University Students’ Mental Health: A Systematic Literature Review Introducing the 4M-Model of Individual-Level Interventions. Front Public Health. 2021;9:699030.

16. Panadero E. A Review of Self-regulated Learning: Six Models and Four Directions for Research. Front Psychol. 2017;8:422.

17. Zimmerman BJ, & Moylan, A. R. Self-regulation: Where metacognition and motivation intersect. In: D. J. Hacker JD, & A. C. Graesser, editor. Handbook of metacognition in education Routledge/Taylor & Francis Group; 2009. p. 299–315.

18. Amir Rad F, Otaki F, Baqain Z, Zary N, Al-Halabi M. Correction: Rapid transition to distance learning due to COVID-19: Perceptions of postgraduate dental learners and instructors. PLoS One. 2021;16(6):e0253683.

19. Otaki F, Zaher S, Du Plessis S, Lakhtakia R, Zary N, Inuwa IM. Introducing the 4Ps Model of Transitioning to Distance Learning: A convergent mixed methods study conducted during the COVID-19 pandemic. PLoS One. 2021;16(7):e0253662.

20. Du Plessis SS, Otaki F, Zaher S, Zary N, Inuwa I, Lakhtakia R. Taking a Leap of Faith: A Study of Abruptly Transitioning an Undergraduate Medical Education Program to Distance-Learning Owing to the COVID-19 Pandemic. JMIR Med Educ. 2021;7(3):e27010.

21. Otaki F, Amir-Rad F, Al-Halabi M, Baqain Z, Zary N. Self-reported adaptability among postgraduate dental learners and their instructors: Accelerated change induced by COVID-19. PLoS One. 2022;17(7):e0270420.

22. Mugford H, O’Connor C, Danelson K, Popoli D. Medical Students’ Perceptions and Retention of Skills From Active Resilience Training. Fam Med. 2022;54(3):213–5.

23. Fenwick TJ. Experiential Learning: A Theoretical Critique from Five Perspectives. Information Series No. 385.. Center on Education and Training for Employment, Columbus, Ohio: ERIC Publications; Opinion Papers; 2001.

24. Quay J. Experience and Participation: Relating Theories of Learning. Journal of Experiental Education. 2016;Volume 26(2).

25. Kolb DA. Experiential Learning: Experience as the Source of Learning and Development. New Jersey: FT Press.; 2014.

26. Gordan S P. Integrating the Experiential Learning Cycle with Educational Supervision. Journal of Educational Supervision. 2022;5(3).

27. Zigmont JJ, Kappus LJ, Sudikoff SN. Theoretical foundations of learning through simulation. Semin Perinatol. 2011;35(2):47–51.

28. Seaman JB M & Quay, J. The Evolution of Experiential Learning Theory: Tracing Lines of Research in the JEE Journal of Experiental Education. 2017;40(4).

29. Gheihman G, Cooper C, Simpkin A. Everyday Resilience: Practical Tools to Promote Resilience Among Medical Students. J Gen Intern Med. 2019;34(4):498–501.

30. Mascarenhas S, Al-Halabi M, Otaki F, Nasaif M, Davis D. Simulation-based education for selected communication skills: exploring the perception of post-graduate dental students. Korean J Med Educ. 2021;33(1):11–25.

31. Liu HB, RE. Focusing on Resilience and Renewal From Stress: The Role of Emotional and Social Intelligence Competencies Frontiers in Psychology 2021;12(685829).

32. Lee CJG. Reconsidering Constructivism in Qualitative Research. Educational Philosophy and Theory. 2012;44(4): 403–12.

33. Yilmaz K. Comparison of Quantitative and Qualitative Research Traditions: epistemological, theoretical, and methodological differences. European Journal of Education, Research, Development and Policy. 2013;48(2):311–25.

34. Cortazzi M. Narrative Analysis. London, UK: Routledge, Taylor & Francis Group; 1993.

35. Cortazzi M, Jin L, Wall D, Cavendish S. Sharing learning through narrative communication. Int J Lang Commun Disord. 2001;36 Suppl:252–7.

36. Braun VC, V Using thematic analysis in psychology. Qualitative Research in Psychology. 2006;3(2):77–101.

37. Kiger ME, Varpio L. Thematic analysis of qualitative data: AMEE Guide No. 131. Med Teach. 2020;42(8):846–54.

38. Seaman J D U., Humberstone, B., Martin, B., Prince, H., and Quay, J. Joint recommendations on reporting empirical research in outdoor, experiential, environmental, and adventure education journals. Journal of Experiential Education 2020;4:348–64.

39. O’Brien BC Hi, Beckman TJ, Reed DA, Cook DA. Standards for reporting qualitative research: a synthesis of recommendations. Academic Medicine. 2014;89(9):1245–51.

40. Levitt Hm BM, Creswell JW, Frost DM, Josselson R, Suarez-Orozco C Standards for qualitative primary, qualitative meta-analytic, and mixed methods research in psychology: The APA Publications and Communications Board task force report. American Psychologist 2018;73(1):26–46.

41. Bandura A. Self-efficacy: toward a unifying theory of behavioral change. Psychol Rev. 1977;84(2):191–215.

42. Howe A, Smajdor A, Stockl A. Towards an understanding of resilience and its relevance to medical training. Med Educ. 2012;46(4):349–56.

43. Goozner M. Health Care: The Causes and Consequences of Slower-Growing Costs Challenge. Taylor & Francis Journals. 2019;62(1):67–76.

44. Miller ED. Loneliness in the Era of COVID-19. Frontiers in Psychology. 2020;11.

45. Harvey M, Coulson, D., & McMaugh, A.. Towards a theory of the Ecology of Reflection: Reflective practice for experiential learning in higher education. Journal of University Teaching & Learning Practice. 2016;13(2).

46. Levine PAB A & Sylvie, J Reintegrating Fragmentation of the Primitive Self: Discussion of “Somatic Experiencing”. Psychoanalytic Dialogues. 2018;28(5): 620–8.

47. Brown KWR, R. & Creswell, J.D Mindfulness: Theoretical Foundations and Evidence for its Salutary Effects. Psychological Inquiry. 2007;18(4):211–37.

48. Lave J, & Wenger, E. Situated learning: Legitimate peripheral participation: Cambridge University Press; 1991.

49. Langdon FJ. Shifting perception and practice: New Zealand beginning teacher induction and mentoring as a pathway to expertise. Journal of Professional Development in Education. 2011;37(2):241–58.

50. Wyatt C, Harper B, Weatherhead S. The experience of group mindfulness-based interventions for individuals with mental health difficulties: a meta-synthesis. Psychother Res. 2014;24(2):214–28.

51. Chambers ES, Bridge MW, Jones DA. Carbohydrate sensing in the human mouth: effects on exercise performance and brain activity. J Physiol. 2009;587(Pt 8):1779–94.

52. Dobkin PL IJ, Amar S.. For whom may participation in a mindfulness-based stress reduction program be contraindicated? Mindfulness. 2012;3(1):44–50.

53. Katz RO, S; Shaw, J & Woodhead, L. The Art of Living in a Digital Age Chicago: The University of Chicago Press; 2021.

54. Hyland T. McDonaldizing Spirituality: Mindfulness, Education, and Consumerism Journal of Transformative Education 2017;15(4).

55. Weinberg A, Creed F. Stress and psychiatric disorder in healthcare professionals and hospital staff. Lancet. 2000;355(9203):533–7.

56. Hassed C. Mindfulness: Is it buddhist or universal? The Humanistic Psychologist. 2021;49(1):72–88

57. Shapiro SL, Schwartz GE, Bonner G. Effects of mindfulness-based stress reduction on medical and premedical students. J Behav Med. 1998;21(6):581–99.

58. Marchand WR. Neural mechanisms of mindfulness and meditation: Evidence from neuroimaging studies. World J Radiol. 2014;6(7):471–9.

59. Danilewitz M, Koszycki D, Maclean H, Sanchez-Campos M, Gonsalves C, Archibald D, et al. Feasibility and effectiveness of an online mindfulness meditation program for medical students. Can Med Educ J. 2018;9(4):e15–e25.

60. Brown KW, Ryan, R.M & Creswell, J.D Mindfulness: Theoretical Foundations and Evidence for its Salutary Effects. Psychological Inquiry. 2007;18(4):211–37.

